# A kinetic model considering the decline of antibody level and vaccination of COVID-19

**DOI:** 10.1101/2021.12.01.21266680

**Authors:** Chuanqing Xu, Xiaotong Huang, Zonghao Zhang, Jingan Cui

## Abstract

In order to evaluate the decline in antibody levels and the impact of vaccination on the spread of the epidemic, we establish COVID-19 dynamic models that consider the decline in antibody levels and the effects of vaccination, and retrospectively evaluate the epidemic situation in England. Based on the epidemic data in England from September 1 to October 31, 2020, considering the continuous decline in the antibody level of COVID-19 recovers, an improved SEIR infectious disease dynamics model that considers the reinfection of recovers due to the decline in antibody levels is established. The kinetic parameters of the SEIR model are obtained by fitting. On this basis, a SEIRV infectious disease dynamic model with vaccination is established to study the impact of different vaccination rates and vaccine failure rates on the development of the epidemic in England. We obtain the lower the vaccine failure rate, the fewer new cases. When the vaccination rateα is fixed at 0.005 (equivalent to 250000 people vaccinated every day), the peak of the epidemic will decrease with the decrease of vaccine failure rate. The peak value when the failure rate is 0.001 is 81.4% lower than the peak value when the failure rate is 0.01, and the peak value when the failure rate is 0.01 is 89.5% lower than the peak value when the failure rate is 0.02. When the failure rate is less than 0.01, the peak time will advance with the decrease of failure rate; when the failure rate is greater than 0.01, the peak time will be delayed with the decrease of failure rate; when the failure rate is 0.01, the peak time is 528 days later than that when the failure rate is 0.001 and 295 days later than that when the failure rate is 0.05. On the 60th day of vaccination, the vaccine failure rate of 0.002 decreases the number of cases by 5.8% compared with the vaccine failure rate of 0.01; on the 70th day of vaccination, the vaccine failure rate of 0.002 reduces the number of cases by 9.1% compared with the vaccine failure rate of 0.01. Therefore, with the extension of time, the vaccine with low failure rate has a more obvious effect on reducing the number of cases than the vaccine with high failure rate. When the vaccine failure rateω is fixed at 0.005, we study the impact of different vaccination rates on the spread of the epidemic in England, the result shows that the peak of epidemic situation decreases with the increase of vaccination rate, and the peak time advance with the increase of vaccination rate, when the vaccination rate is 0.025, the peak decreases by 74.8% and the peak time was 114 days earlier than that when the vaccination rate is 0.005. Therefore, the higher the vaccine efficiency and vaccination rate, the lower the peak of the epidemic. On the basis of improving the effectiveness of vaccines, increasing the vaccination rate is of practical significance for controlling the spread of the epidemic.

## Introduction

It has been almost two years since the outbreak of COVID-19. The research in the world has given us a certain understanding of the virus. This is a serious respiratory disease caused by coronavirus^[1,2]^, The International Committee on Taxonomy of viruses named the virus SARS-CoV-2 is mainly transmitted through respiratory tract. The clinical symptoms of infected people are mostly fever, dry cough and fatigue, and some patients are asymptomatic infected people. When the disease is serious, it will also cause organ failure and even death^[3,4]^.

At present, the pandemic of covid-19 is still in progress. According to the latest WHO epidemic situation, more than 70 million new cases have been diagnosed in Europe by the end of October 2021^[5]^. Britain is one of the country most seriously affected by the epidemic in Europe, with more than 8 million cumulative cases, of which more than 7 million cases have been confirmed in England alone^[6]^, accounting for 85% of the total number of cumulative reports in Britain. Since more than 80% of confirmed cases in Britain are concentrated in England, we intend to do some theoretical research based on the data of England for the reference of government decision-making. Figure 1 shows the daily new cases data of COVID-19 epidemic in England during 2020. At the beginning of March, due to the obvious increase of new cases, the British government began to implement the first large-scale social blockade at the end of March and gradually unsealed at the beginning of June. This phase corresponds to the phase in which the first small peak appears in Figure 1.In the three months after the first unsealing, the epidemic situation in the region had been relatively gentle. However, since September, the number of new cases per day had increased significantly. By the end of October, the number of new cases per day had reached about 20000.At this time, the British government once again announced that it would implement the second large-scale social blockade on November 5, which lasted until December 2^nd^. Soon after the second unsealing, the third wave of more large-scale epidemic began, and there were reports that a mutant virus with stronger transmission ability was detected, and the epidemic reached a stage that was difficult to control. On December 20^th^, Britain ushered in the third social blockade, which lasted until February 2021.

**Fig.1.**
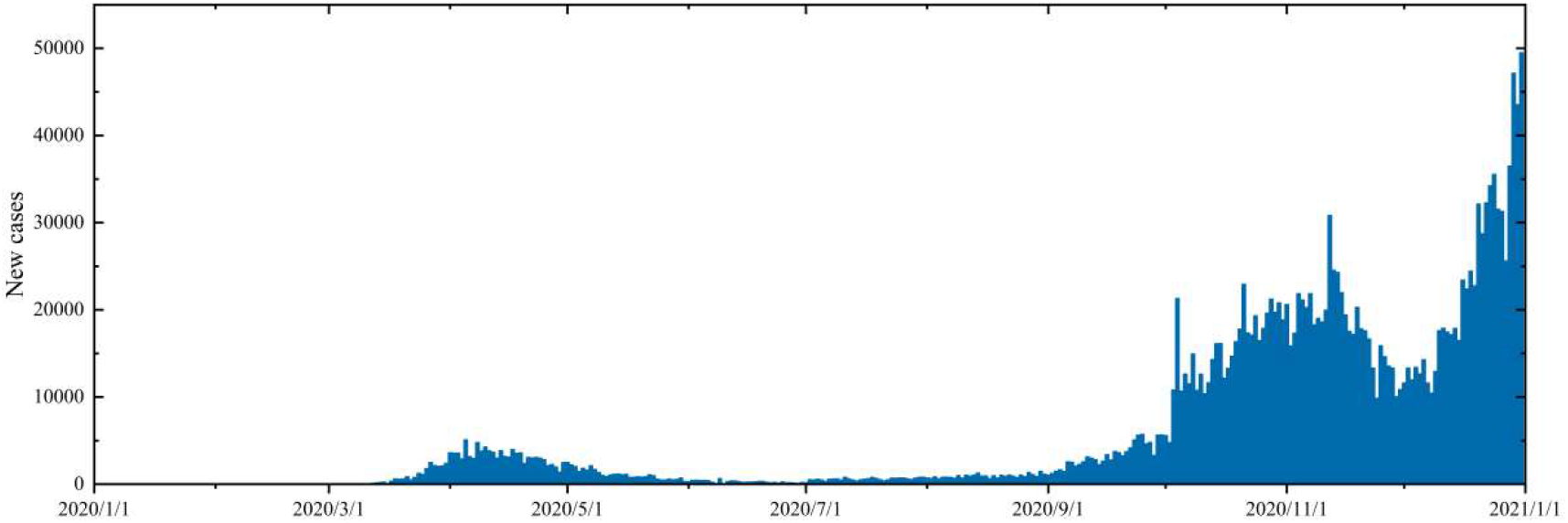
Daily new case data of COVID-19 in England in 2020

The results of an antibody level detection study based on 365000 people by H.ward et al showed that only 4.4% of adults detected IgG antibodies using LFIA at the beginning of the second wave of infection in England^[7]^.A study on the antibody positive rate of asymptomatic and symptomatic infected people in Wanzhou District of Chongqing by long Quanxin et al showed that after 8 weeks of infection, the antibody concentration of more than 90% of the participants decreased by more than 70%^[8]^.A study on the neutralizing antibody level of SARS-CoV-2 patients within three months after infection found that after analyzing the serum samples of 65 SARS-CoV-2 infected patients, 95% of the cases had seroconversion^[9]^.Some researchers found that the antibody titers of the two virus strains decreased significantly, and the delta variant virus strain decreased more significantly than the wild strain^[10-12]^.

Since the antibody level in the convalescent population of COVID-19 will decrease over time, the recovered individuals will get the chance to infect the virus again. In the first model, we consider that a certain proportion of restorers will become susceptible again, so as to explore the impact of the decline of antibody level on the transmission of COVID-19 epidemic in England. SEIR (susceptible exposed infectious recovered) mathematical model is a key tool for studying the spread of infectious diseases. It can clearly describe the dynamic relationship between warehouses during virus transmission and give a relatively accurate prediction for the development trend of infectious diseases. Wintachai. P ^[13]^ et al predicted the change trend of COVID-19 cases over time by establishing SEIR infectious disease dynamics model considering different vaccination rates. A. Fuady et al considered the different vaccination time to establish the dynamics model of infectious diseases, so as to explore the inhibitory effect of the change of vaccination time on the spread of covid-19 epidemic ^[14]^.

This study selects the daily new data of COVID-19 epidemic in England from September 1 to October 31, 2020, that is, the data during the second wave of epidemic growth in England. The reason is that the British government adopted natural immunization measures without any intervention policy during this period, so we can only consider the impact of the decline of antibody level on the transmission of new crown in the first model.

## 1. Improved SEIR model for the decline of antibody level in population

### 1.1 Model

The SEIR infectious disease dynamics model considering the decline of antibody level in the restorer is shown in Figure 2. We divide the total population *N* into four categories: susceptible people *S*, exposed people *E*, infected people I and recovered people *R*. among *β* indicates the rate of disease transmission, *σ* indicates the conversion rate from exposed people to infected people, *γ* represents the disease recovery rate, *μ* represents the mortality due to disease, and *P* represents the conversion rate from recovered people to susceptible people. The propagation mechanism of the model is shown in the figure below:

**Fig.2.**
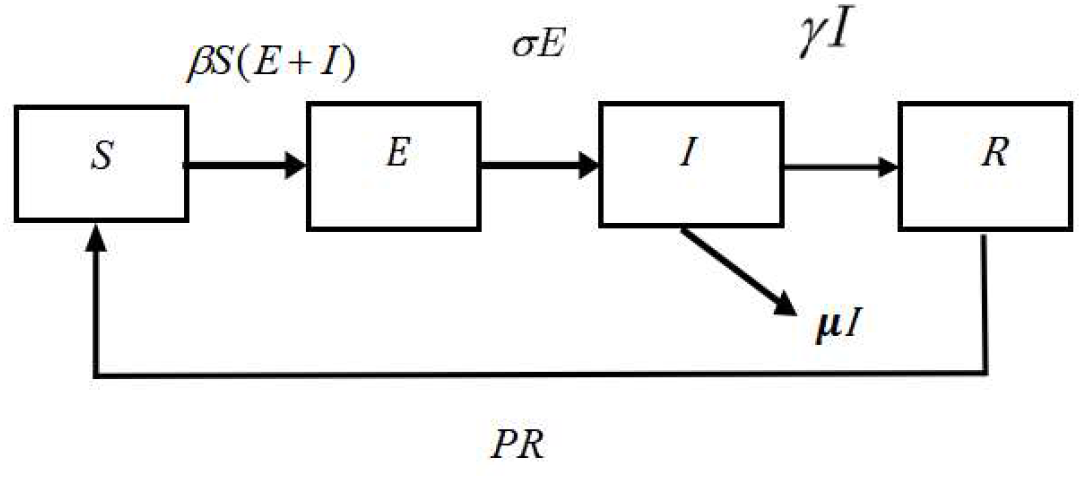
COVID-19 transmission considering decreased antibody levels

According to the above COVID-19 transmission diagram, we establish the infectious disease dynamics model as follows:

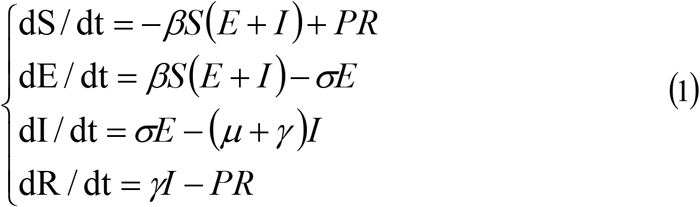

### 1.2 Model analysis

#### 1.2.1 Equilibrium point of model

Let

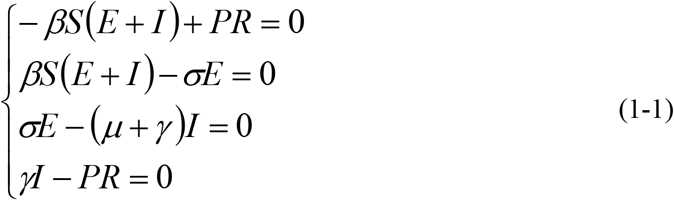

When *I* = 0, the disease-free equilibrium point of model (1) is *E*_01_ = (*S*_0_, 0,0,0). When *I* ≠ 0, by solving equations (1-1):

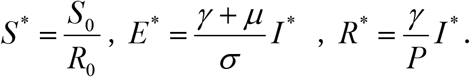

Therefore, the endemic equilibrium point of model (1) is:

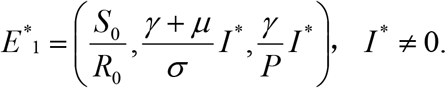

#### 1.2.2 Basic regeneration number

The basic regeneration number refers to the average number of secondary cases after an infected individual enters the susceptible population^[15]^,the next generation matrix is the most commonly used method to calculate the basic regeneration number.

In order to calculate the basic regeneration number of model (1), we take:

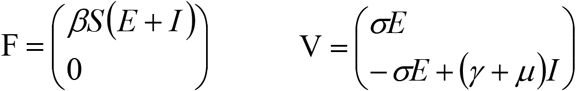

The Jacobian matrices of F and V are:

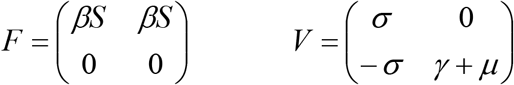

At disease-free equilibrium:

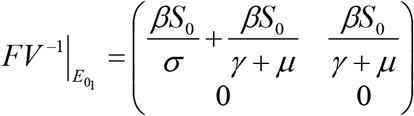

The basic regeneration number *R*_0_ is the spectral radius of 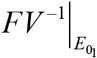, so the expression is

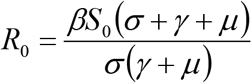

#### 1.2.3 Stability analysis of disease-free equilibrium

##### Theorem 1.2.3

When *R*_0_ <1, the model (1) is stable at the disease-free equilibrium point, otherwise it is unstable.

**Proof**. The Jacobian matrix of model (1) at *E*_01_is:

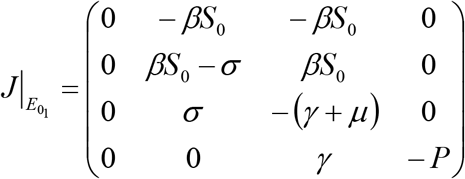

The matrix eigenvalue satisfies the following formula:

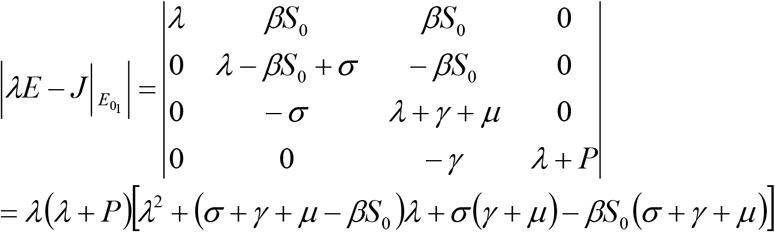

Obviously *λ*_1_ =0, *λ*_2_ = -*P* <0, the other two eigenvalues *λ*_3_, *λ*_4_ satisfy the equation *P*(*λ*) = *λ*^2^ + *a*_1_*λ* + *a*_2_ = 0

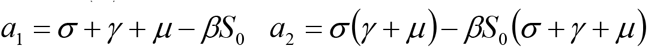

Easy to know 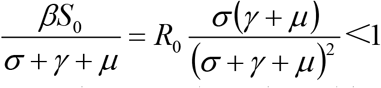, therefore, *a*_1_ = *σ* + *γ* + *μ* − *βS*_0_>0

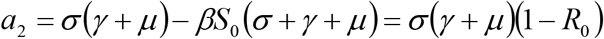

So, when 0<*R*_0_<1 *a*_1_>0, *a*_2_>0.

According to Hurwitz discriminant method, when 0<*R*_0_<1, the characteristic roots *λ*_3_, *λ*_4_ of *P*(*λ*) have negative real parts. Therefore *λ*_3_<0, *λ*_4_<0

To sum up, among the eigenvalues of the Jacobian matrix of model (1), four have negative real parts and one is zero, so model (1) is stable at the disease-free equilibrium point, otherwise it is unstable.

### 1.3 Numerical simulation

The results of numerical simulation of new cases in England from September 1 to October 31 by nonlinear least square method are shown in Figure 3.Within the acceptable error range of population scale, we set the initial value of susceptible persons as *S*_0_ = 55699400, that is, the total population of England, the initial value of exposed persons as *E*_0_ = 6500, the initial value of infected persons as *I*_0_ = 1041, that is, the number of new cases in England on September 1, and the initial value of recovered person as 200.In the model, the conversion rate from exposed person to infected person *σ* = 1/12, the recovery rate of disease *γ* = 1/14 and the mortality rate due to disease *μ* = 0.003 are selected according to the actual transmission of infectious diseases, and the disease transmission rate *β* = 1.1295×10^−9^ and the conversion rate from recovered person to susceptible person *P* = 1.8888×10^−6^ are fitted.

**Fig.3.**
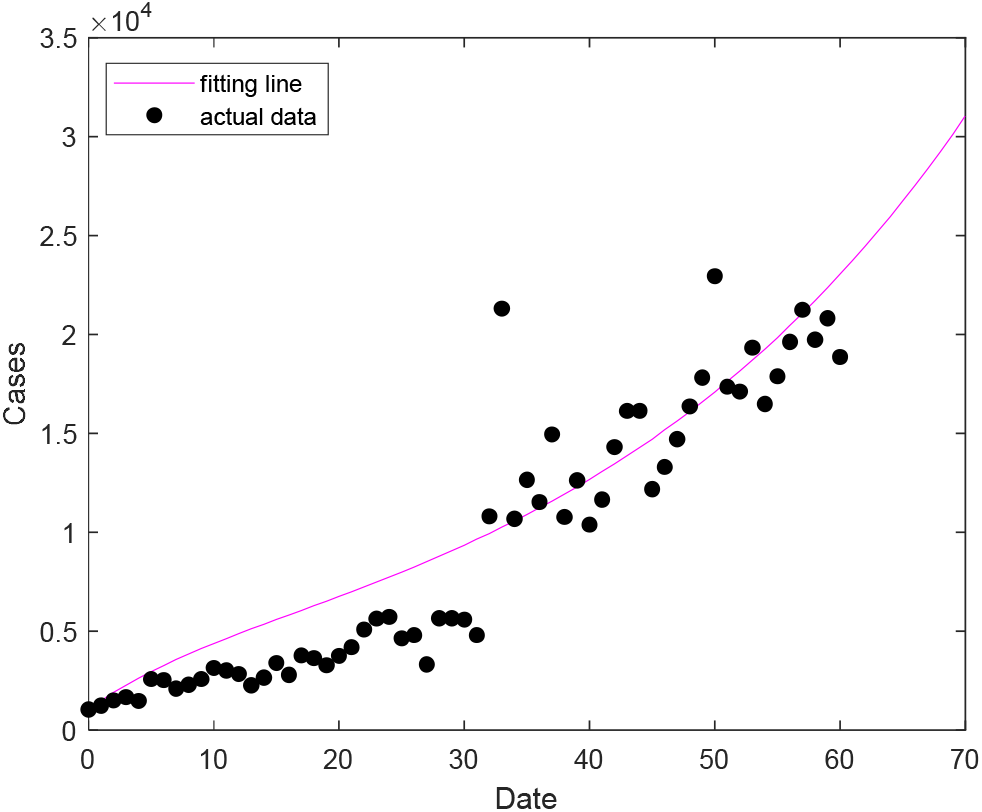
Data simulation in England

The fitting results of the improved SEIR epidemic dynamics model are in good agreement with the data of England, and can well reflect the growth trend of COVID-19 epidemic in England.

## 2. SEIRV model of vaccination

### 2.1 The model

The antibody level will decline over time, and the method of relying on natural immunity to obtain group immunity becomes infeasible. Therefore, on the basis of the above model (1), we add the vaccinated warehouse *V* (the successfully vaccinated population) to establish the SEIRV model. In this model, we consider vaccinating the susceptible population at a fixed vaccination rate every day, and explore the impact of different injection rates and failure rates on the epidemic situation in England. Where *α* represents the daily vaccination rate of vaccine and *ω* represents the vaccine failure rate. The meaning of other parameters in the model is the same as that in model (1). The propagation mechanism of the model is shown in the Fig.4 below:

**Fig.4.**
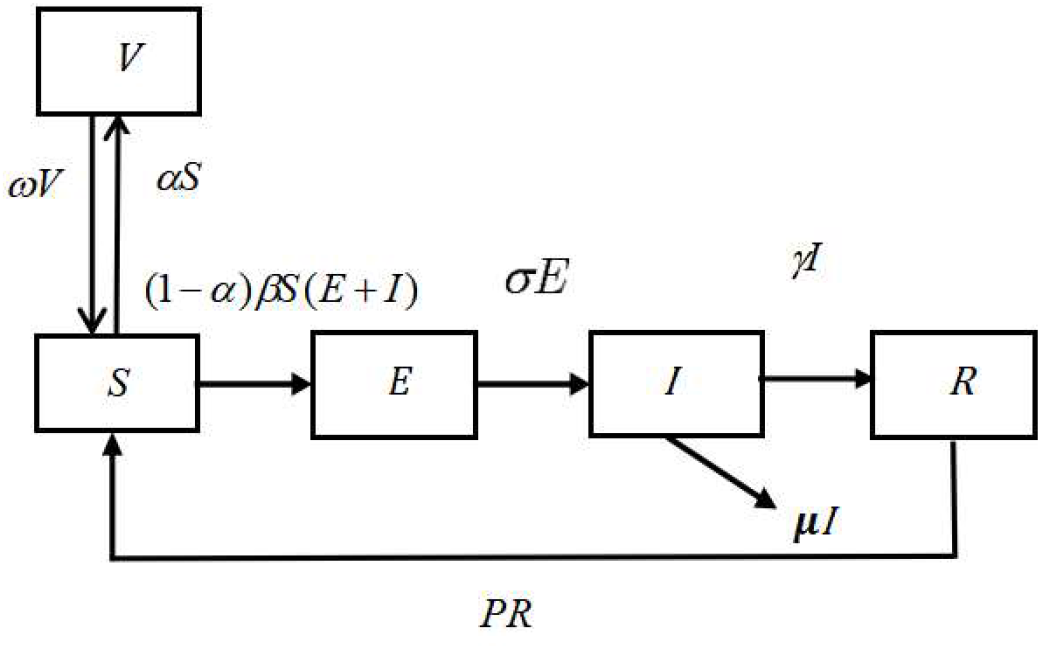
COVID-19 transmission considering decreased antibody levels and vaccination

According to the above COVID-19 transmission diagram, the infectious disease dynamics model we establish is:

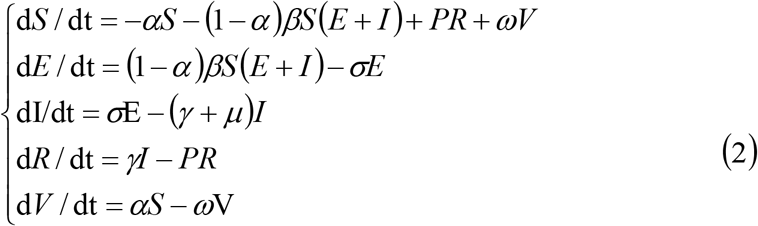

### 2.2 Model analysis

#### 2.2.1 Equilibrium point of model

Model (2) is transformed into a system of equations:

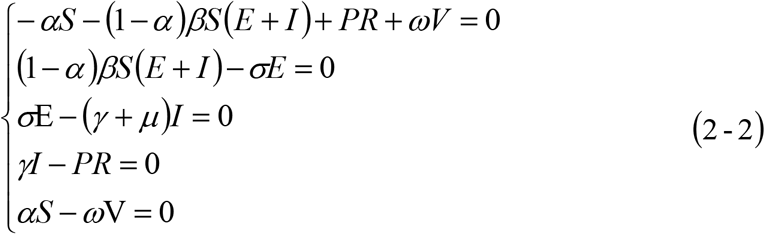

When *I* = 0, there are:

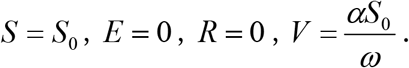

That is, the disease-free equilibrium point of model 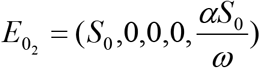

When *I* ≠ 0, by solving equations (2-2):

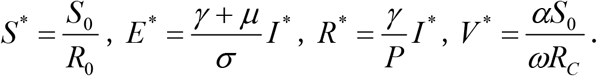

Therefore, the endemic equilibrium point of model (2) is:

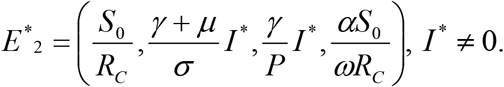

#### 2.2.2 Control regeneration number

The number of controlled regeneration refers to the number of people who can be infected by an infected person during the infection period under certain prevention and control measures^[16]^. It is also calculated by the method of next generation matrix.

In order to calculate the control regeneration number of model (2), we take:

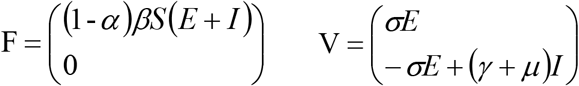

The Jacobian matrices of F and V are:

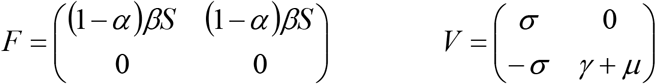

At disease-free equilibrium:

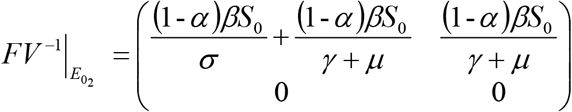

The control regeneration number *R*_0_ is the spectral radius of 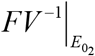, and the expression is:

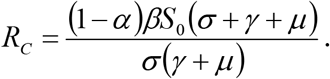

#### 2.2.3 Stability analysis of disease-free equilibrium

##### Theorem 2.2.3

When *R*_*C*_ <1, the model (2) is locally asymptotically stable at the disease-free equilibrium point, otherwise it is unstable.

**Proof**. The Jacobian matrix of model (3) at 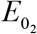 is:

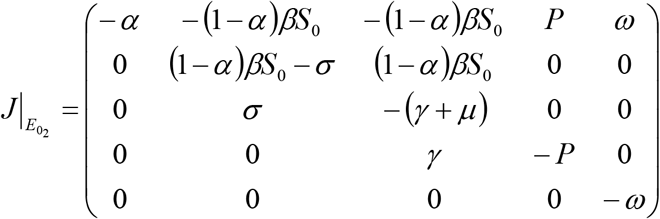

The matrix eigenvalue satisfies the following formula:

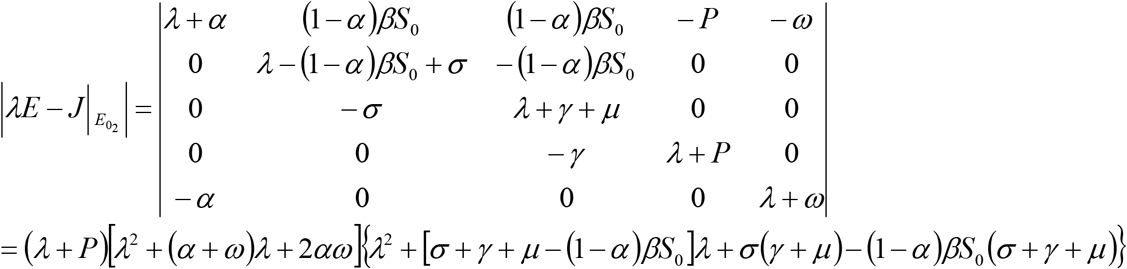

Obviously *λ*_1_ = -*P* <0, the other two eigenvalues *λ*_2_, *λ*_3_ satisfy the equation

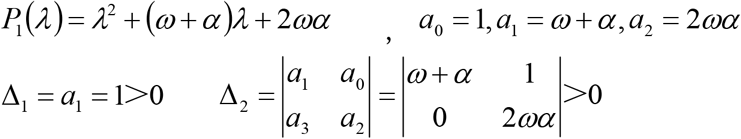

It can be seen from Hurwitz discriminant that *λ*_2_, *λ*_3_ all have negative real parts. *λ*_4_ and *λ*_5_ satisfy the equation *P*_2_(*λ*) = *λ*^2^ + *B*_1_ *λ* + *B*_2_

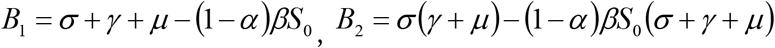

It is the same as the proof of theorem 1.2.3:

Similarly, it can be seen from Hurwitz discriminant that *λ*_4_, *λ*_5_ all have negative real parts, when 0<*R*_*C*_<1 *B*_1_>0, *B*_2_>0.

To sum up, all eigenvalues of the Jacobian matrix of model (2) have negative real parts, so model (2) is locally asymptotically stable at the disease-free equilibrium point, otherwise it is unstable.

### 2.3 Numerical simulation

In the SEIRV model, *α* refers to the daily vaccination rate, which is further explained here. The overall immunization rates corresponding to different daily vaccination rates (60 days after vaccination) is shown in Fig.5. The total population of England is about 50 million, *α* = 0.005 means that 250000 people are vaccinated every day. After 60 days of continuous vaccination, the overall immunization rate is 25.9%.

**Fig.5.**
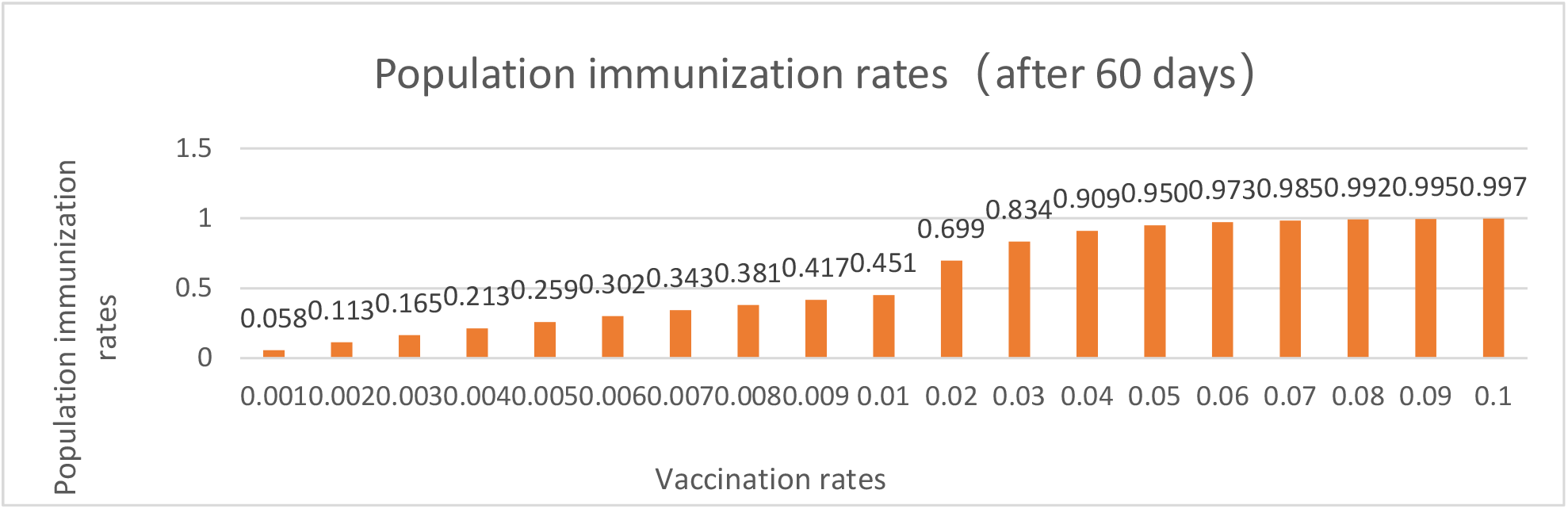
The overall immunization rates of the population after 60 days of continuous vaccination with different vaccination rates

In SEIRV model, the meaning of vaccine failure rate *ω* refers to the reciprocal of the duration of vaccine effectiveness, that is, each corresponds to a duration of vaccine effectiveness. The data details are shown in Table 1.

**Table1.** Vaccine failure rate and duration of vaccine effectiveness

| Vaccine failure rates | Duration of vaccine effectiveness |
| --- | --- |
| 0 | FOREVER |
| 0.001 | 1000 |
| 0.002 | 500 |
| 0.003 | 333 |
| 0.004 | 250 |
| 0.005 | 200 |
| 0.006 | 167 |
| 0.007 | 143 |
| 0.008 | 125 |
| 0.009 | 111 |
| 0.01 | 100 |
| 0.02 | 50 |
| 0.03 | 33 |
| 0.04 | 25 |
| 0.05 | 20 |

#### 2.3.1 Effects of different vaccine failure rates on the epidemic situation in England

In order to evaluate the role of vaccines in epidemic control, we investigate the impact of different vaccination rates and failure rates on the spread of COVID-19 epidemic in England. The prediction results are shown in the figure below. We change the vaccine failure rate when the fixed vaccination rate is 0.005, and simulate the impact of different failure rates on the spread of COVID-19 epidemic in England. The results are shown in Figure 6-8.In Figure 6, the failure rate is taken from 0.001 to 0.01 in steps of 0.001; in Fig. 7, the failure rate is taken from 0.01 to 0.05 in steps of 0.01; in Figure 8, the failure rate is taken from 0.01 to 0.02 in steps of 0.001. As can be seen from figure 6-8, with the decrease of vaccine failure rate, the peak of epidemic situation will gradually decrease. When the failure rate is less than 0.01, the peak time will advance with the decrease of failure rate; when the failure rate is greater than 0.01, the peak time will be delayed with the decrease of failure rate. If only figures 6 and 7 are used to illustrate that 0.01 is the boundary value of peak time trend change, it may be coincidental because the failure rate less than 0.01 is taken as the step of 0.001, and the failure rate greater than 0.01 is taken as the step of 0.01. Therefore, we further take the failure rate from 0.01 to 0.02 in steps of 0.001 for simulation, as shown in Figure 8. It can be seen from figure 8 that the time of peak value is delayed with the decrease of failure efficiency. In general, when the step of failure rate change is 0.001, the simulation shows that when the fixed vaccination rate is 0.005, the peak of epidemic situation will decrease with the decrease of failure efficiency. When the failure rate is less than 0.01, the peak time will advance with the decrease of failure efficiency; when the failure rate is greater than 0.01, the peak time will be delayed with the decrease of failure efficiency.

**Fig.6.**
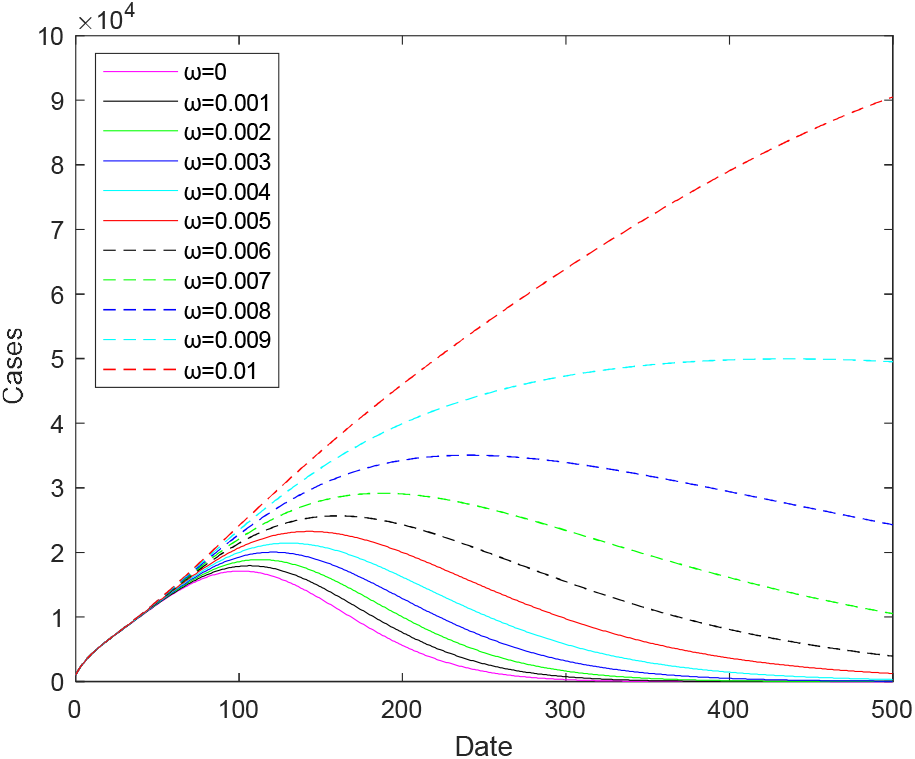
Prediction of COVID-19 epidemic in England with different vaccine failure rates when the vaccination rate is 0.005

**Fig.7.**
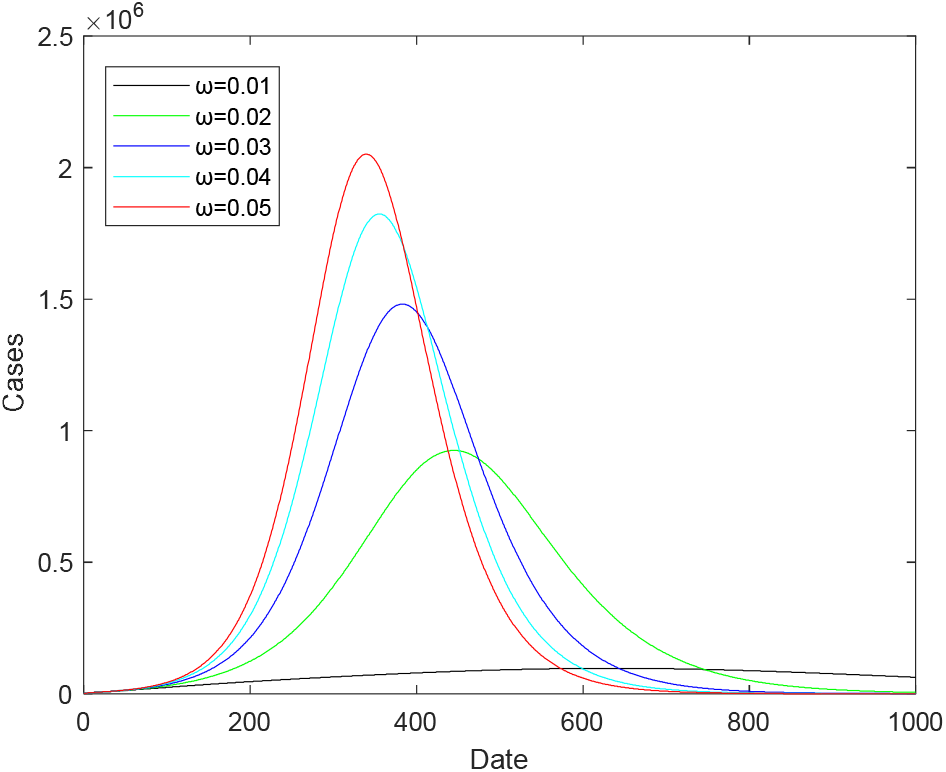
Prediction of COVID-19 epidemic in England with different vaccine failure rates when the vaccination rate is 0.005

**Fig.8.**
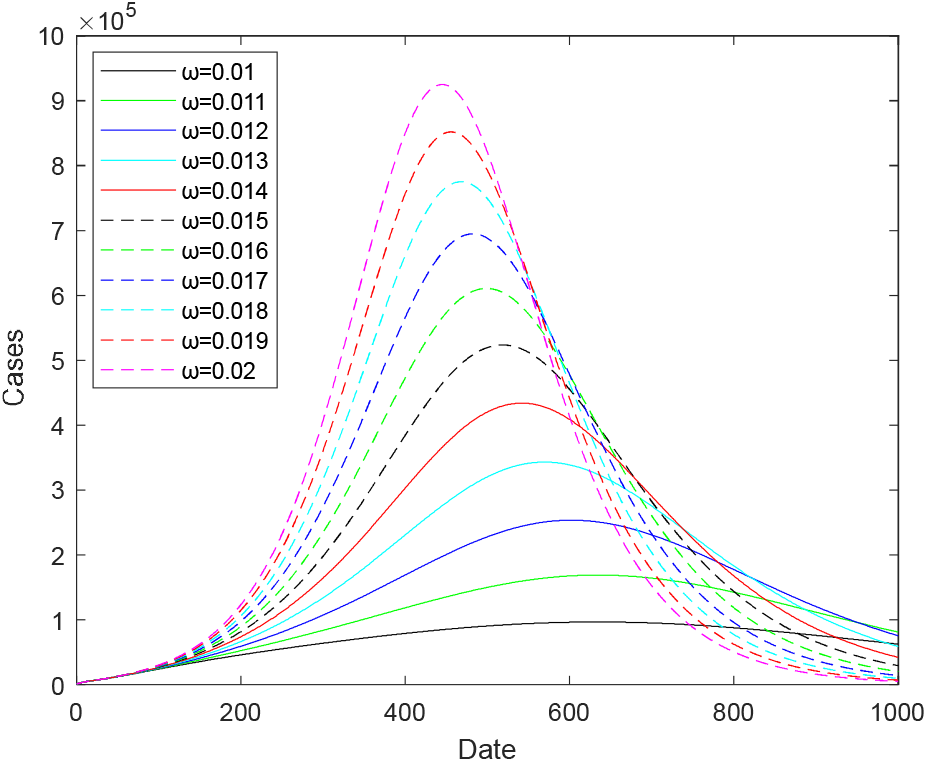
Prediction of COVID-19 epidemic in England with different vaccine failure rates when the vaccination rate is 0.005

When the vaccination rate is 0.005, the peak histogram of epidemic situation under different vaccine failure rates is shown in Fig.9 and Fig.10. When the value of failure rate is large (greater than 0.02), the peak will increase significantly (Fig. 10). The peak value when the failure rate is 0.001 is 81.4% lower than the peak value when the failure rate is 0.01, and the peak value when the failure rate is 0.01 is 89.5% lower than the peak value when the failure rate is 0.02. Therefore, the lower the vaccine failure rate, the lower the peak of the epidemic. The peak values under different failure rates are shown in Table 2.

**Table2.** when the vaccination rate is 0.005, the peak value, peak time and days of vaccine effectiveness of different vaccine failure rates

| Vaccine failure rates | Duration of vaccine effectiveness | Peak value of cases | Peak time (days) |
| --- | --- | --- | --- |
| 0 | FOREVER | 17126 | 101 |
| 0.001 | 1000 | 17934 | 106 |
| 0.002 | 500 | 18887 | 113 |
| 0.003 | 333 | 20032 | 121 |
| 0.004 | 250 | 21443 | 130 |
| 0.005 | 200 | 23244 | 143 |
| 0.006 | 167 | 25658 | 160 |
| 0.007 | 143 | 29159 | 187 |
| 0.008 | 125 | 35046 | 239 |
| 0.009 | 111 | 49967 | 435 |
| 0.01 | 100 | 96669 | 634 |
| 0.02 | 50 | 924980 | 445 |
| 0.03 | 33 | 1481000 | 383 |
| 0.04 | 25 | 1823700 | 355 |
| 0.05 | 20 | 2050800 | 339 |

**Fig.9.**
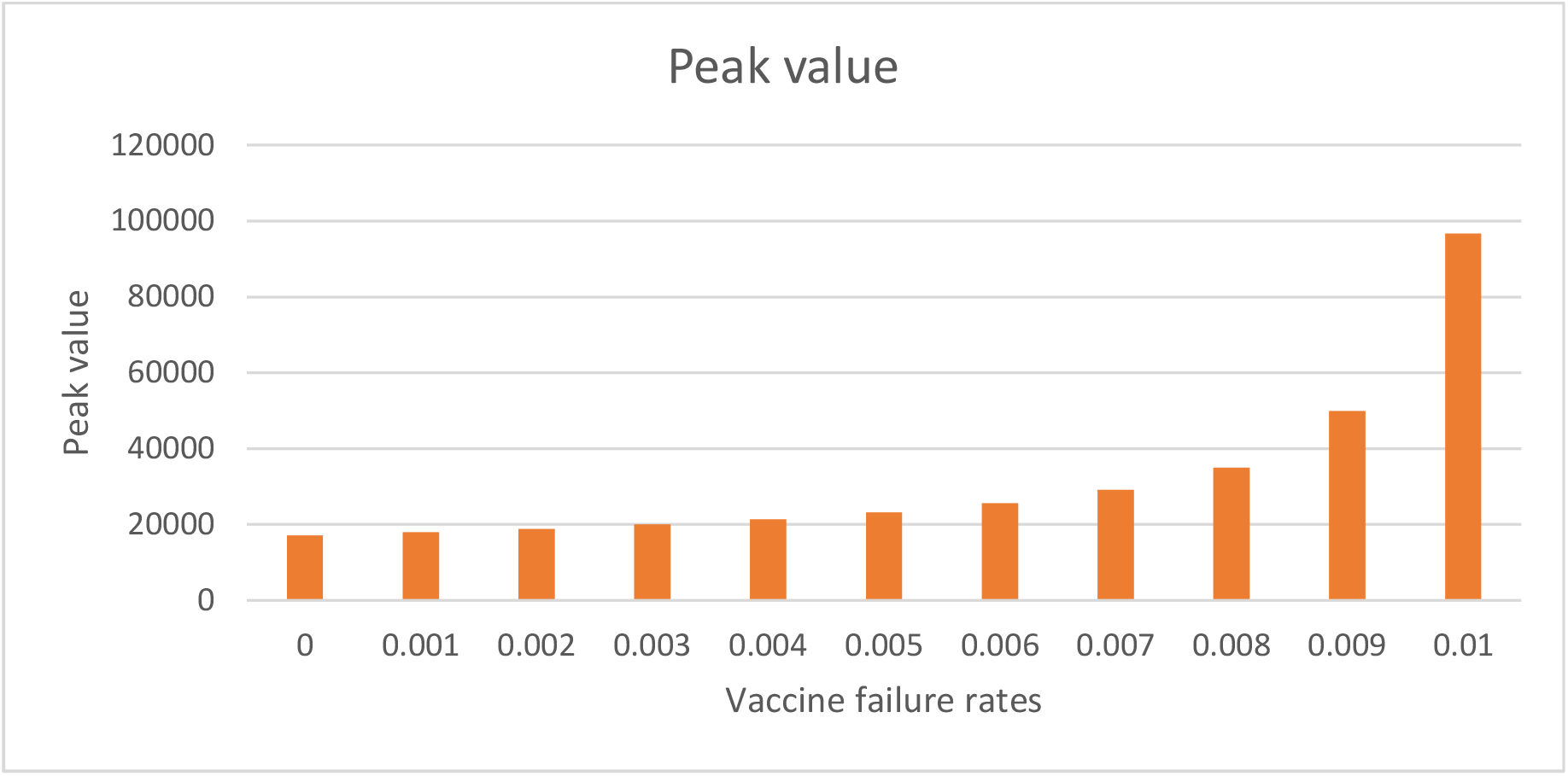
The peak of cases in England with different vaccine failure rates when the vaccination rate is 0.005

**Fig.10.**
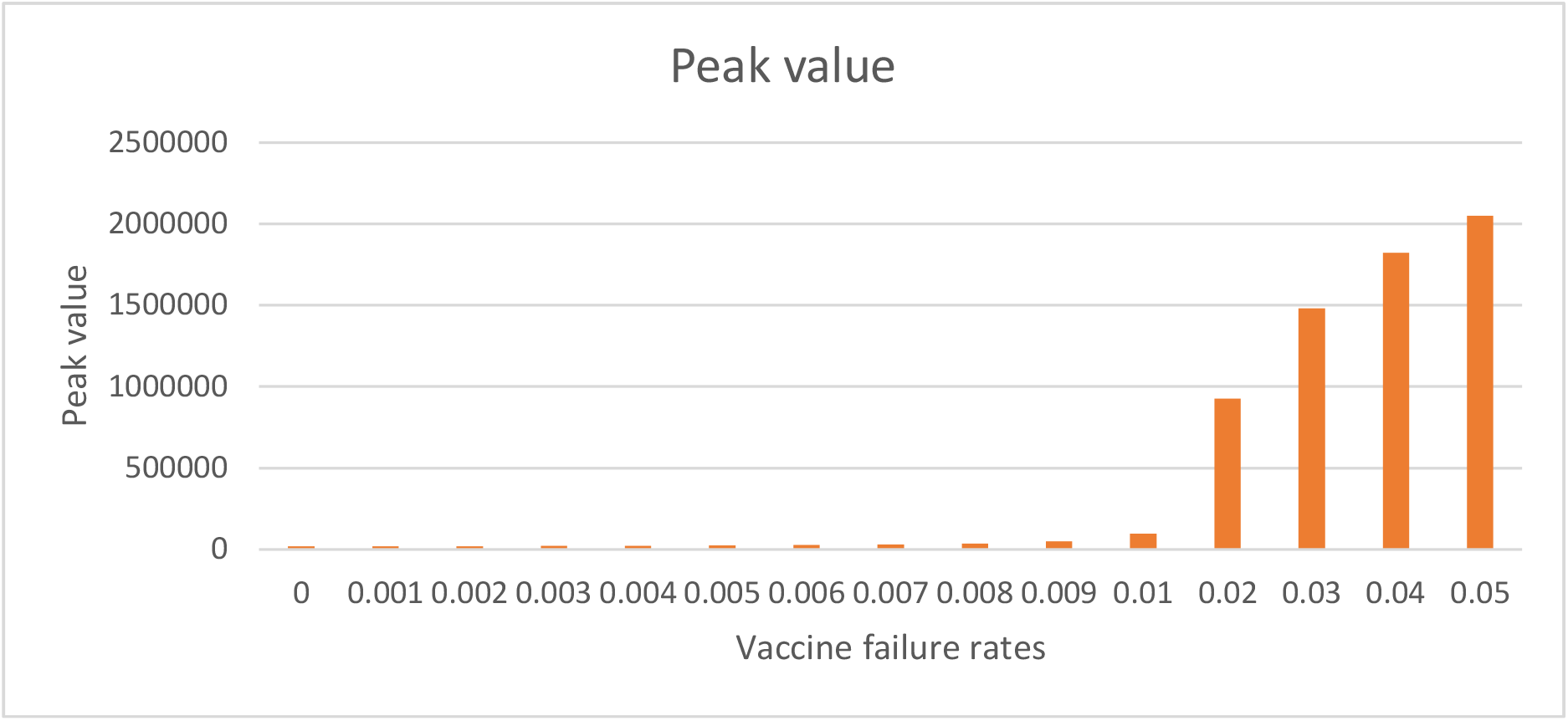
The peak of cases in England with different vaccine failure rates when the vaccination rate is 0.005

When the vaccination rate is 0.005, the histogram of peak time of epidemic situation under different vaccine failure rates is shown in Fig. 11. It can be seen when the failure rate is less than 0.01, the peak time will advance with the decrease of failure rate; when the failure rate is greater than 0.01, the peak time will be delayed with the decrease of failure rate. When the failure rate is 0.01, the peak time is 528 days later than that when the failure rate is 0.001 and 295 days later than that when the failure rate is 0.05. The peak time under different failure rates is shown in Table 2.

**Fig.11.**
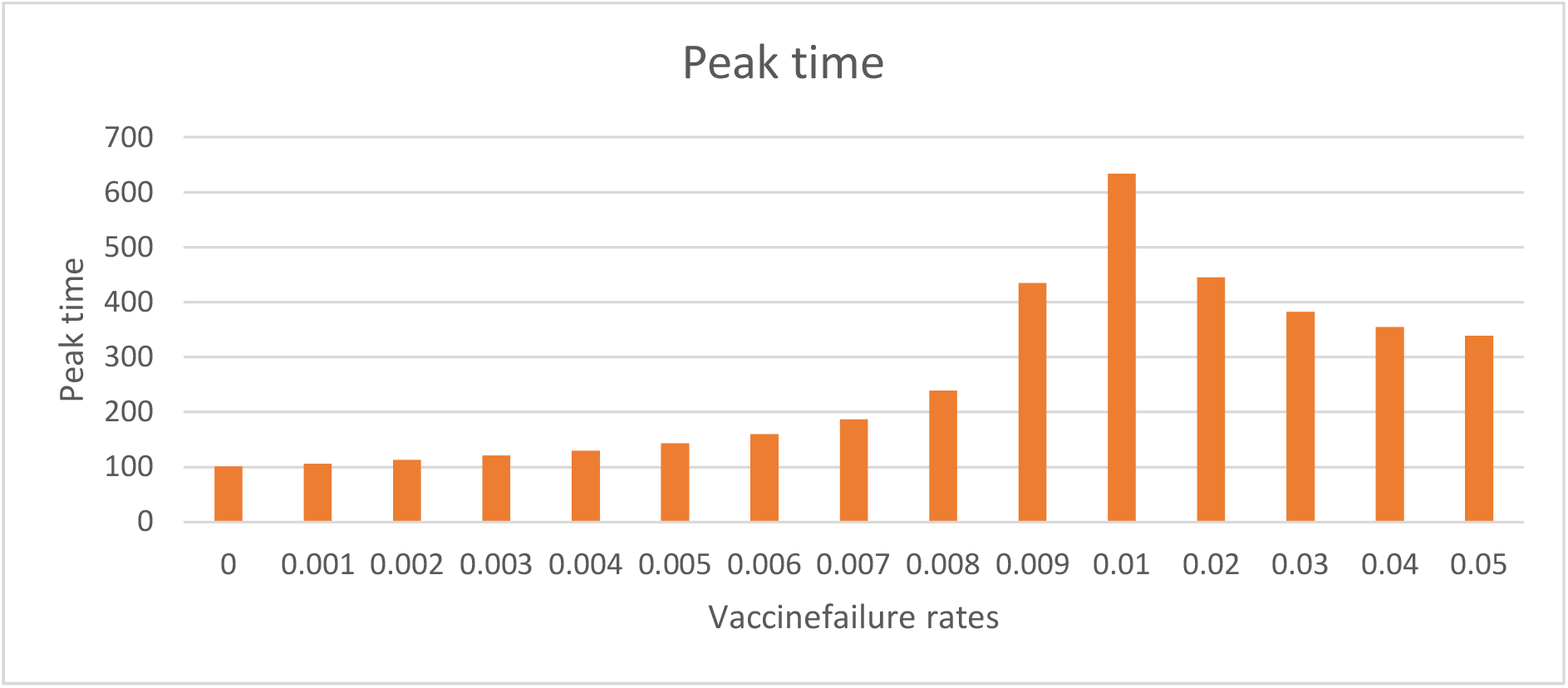
The peak time of cases in England with different vaccine rates when the vaccination rate is 0.005

We also discuss the number of new cases per day at 30, 40, 50, 60 and 70 days in England with different failure rates when the vaccination rate is 0.005. The results are shown in Fig.12. It can be seen from the figure that at the same time, the smaller the failure rate, the fewer the number of cases. On the 60th day of vaccination, the vaccine with failure rate of 0.002 is 5.8% lower than the vaccine with failure rate of 0.01; On the 70th day of vaccination, the vaccine with failure rate of 0.002 is 9.1% lower than the vaccine with failure rate of 0.01. Therefore, with the extension of time, the vaccine with low failure rate is more effective in reducing the number of cases than the vaccine with high failure rate. The number of cases is shown in Table 3.

**Fig.12.**
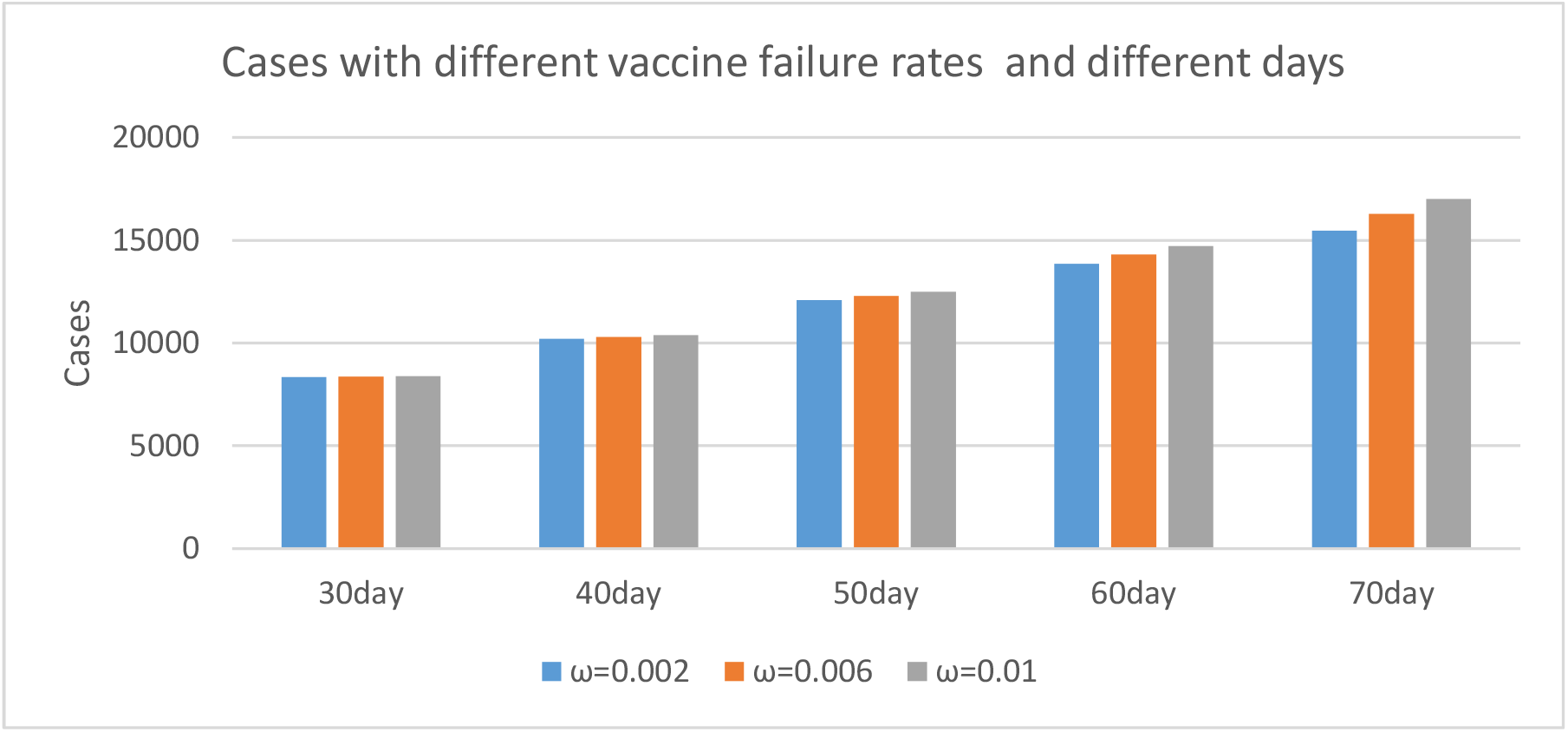
Cases with different vaccine failure rates at 30, 40, 50, 60 and 70 days in England when the vaccination rate is 0.005

**Table 3.** Cases with different vaccine failure rates at 30, 40, 50, 60 and 70 days in England when the vaccination rate is 0.005

|  | 30day | 40day | 50day | 60day | 70day |
| --- | --- | --- | --- | --- | --- |
| $\omega=0.002$ | 8337 | 10214 | 12079 | 13862 | 15476 |
| $\omega=0.006$ | 8366 | 10305 | 12298 | 14307 | 16276 |
| $\omega=0.01$ | 8393 | 10390 | 12502 | 14720 | 17020 |

When the vaccination rate is 0.01, we discuss the impact of different vaccine failure rates on the spread of the epidemic in England, the results are shown in Fig.13-16. The simulation results show that the smaller the failure rate is, the lower the peak of epidemic situation is, and 0.019 is the boundary value of failure rate. When the failure rate is less than 0.019, the peak time will advance with the decrease of failure rate; when the failure rate is greater than 0.019, the peak time will be delayed with the decrease of failure rate. Some data are shown in Table 4.

**Fig.13.**
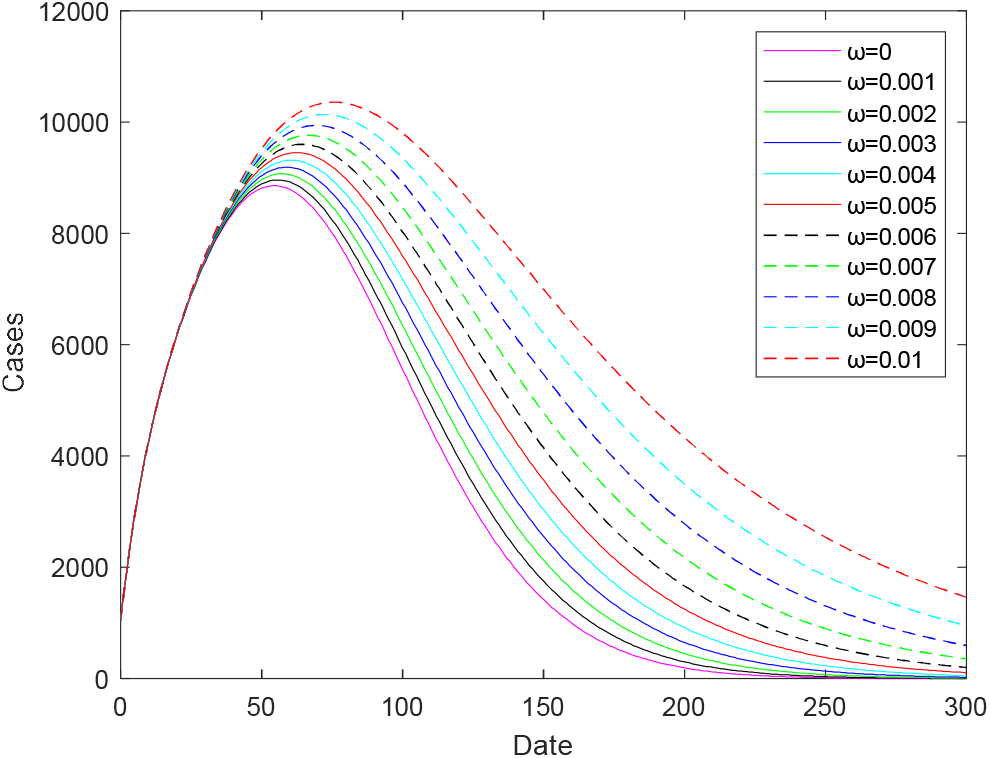
Prediction of COVID-19 epidemic in England with different vaccine failure rates when the vaccination rate is 0.01

**Fig.14.**
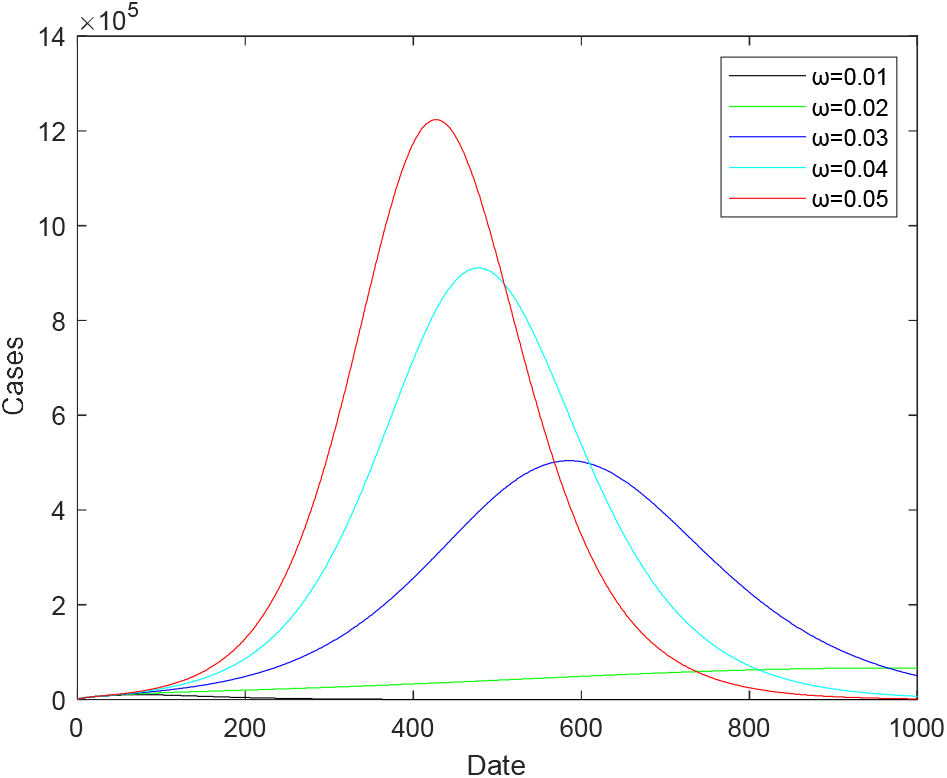
Prediction of COVID-19 epidemic in England with different vaccine failure rates when the vaccination rate is 0.01

**Fig.15.**
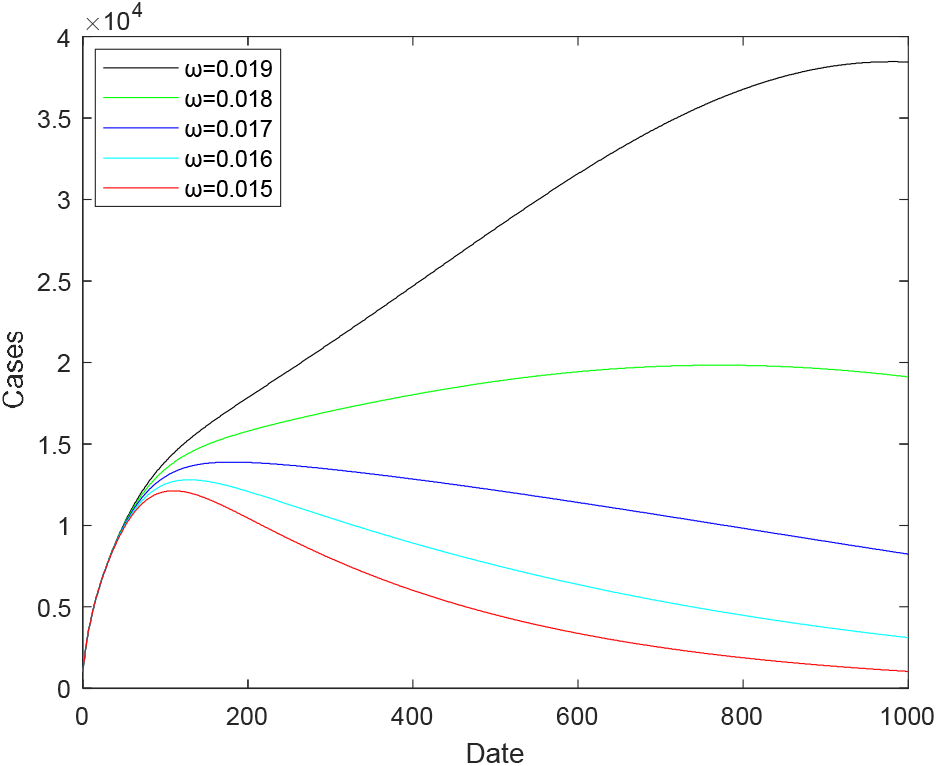
Prediction of COVID-19 epidemic in England with different vaccine failure rates when the vaccination rate is 0.01

**Fig.16.**
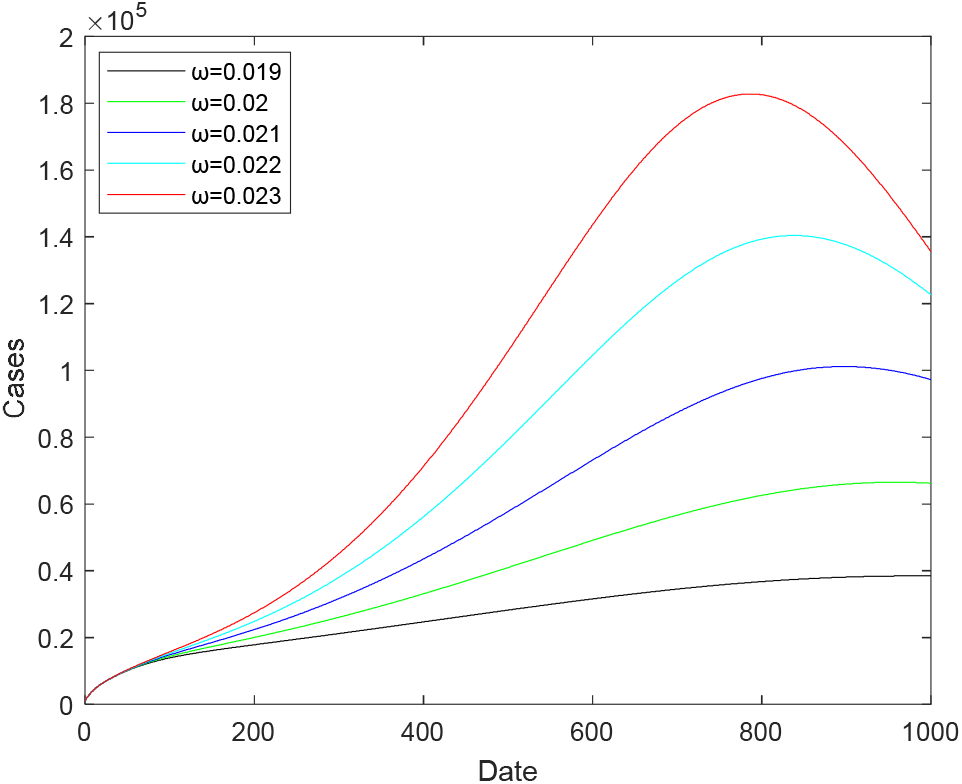
Prediction of COVID-19 epidemic in England with different vaccine failure Table4 When the vaccination rate is 0.01, the peak value and peak time of different vaccine failure

**Table4.**
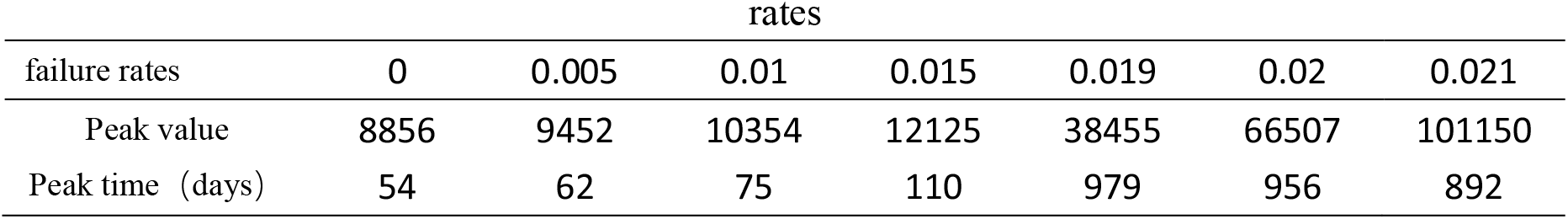
When the vaccination rate is 0.01, the peak value and peak time of different vaccine failure

#### 2.3.2 Effects of different vaccination rates on the epidemic situation in England

When the vaccine failure rate is 0.005, the impact of different vaccination rates on the spread of the epidemic in England is shown in Fig.17. The vaccination rates change from 0.005 to 0.025 in steps of 0.005. According to the figure, the peak of epidemic situation decreases with the increase of vaccination rate, and the peak time advances with the increase of vaccination rate. When the vaccination rate is 0.025, the peak value decrease by 74.8% and the peak time is 114 days earlier than that when the vaccination rate is 0.005. The result is given in table 4.

**Fig.17.**
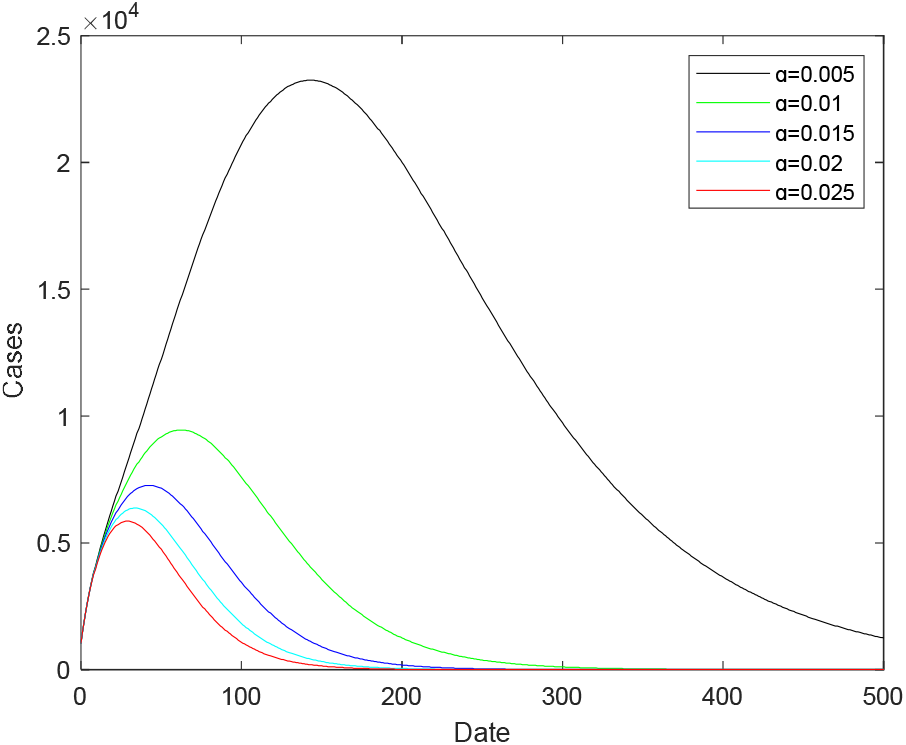
Prediction of different vaccination rates for cases in England when the vaccine failure rate is 0.005

**Table4.** When the vaccine failure rates is 0.005, the peak value and peak time of different vaccination rate

| Vaccination rate | 0.005 | 0.01 | 0.015 | 0.02 | 0.025 |
| --- | --- | --- | --- | --- | --- |
| Peak value | 23244 | 9452 | 7269 | 6370 | 5857 |
| Peak time (days) | 143 | 62 | 42 | 34 | 29 |

## 3 Conclusion

The first part of this study is to establish an improved SEIR model of COVID-19 transmission in England on the basis of considering the decline of antibody levels in restorers. In the second part, based on the decline of antibody level, considering the vaccination of susceptible people at a fixed vaccination rate every day, the SEIRV model of vaccination is established. We calculate the basic regeneration number of model (1) and the control regeneration number of model (2), calculate the disease-free equilibrium point and endemic equilibrium point of the two models, and analyze the stability of the model.

In the numerical simulation part, we further explain the vaccination rate and failure rate: *α* =0.005 means that 250000 people are vaccinated every day on the basis of 50 million people in England, on the basis of this vaccination rate, the overall immunization rate was 25.9% after 60 days of continuous vaccination. In addition, the meaning of vaccine failure rate *ω* refers to the reciprocal of the duration of vaccine effectiveness, that is, each corresponds to a duration of vaccine effectiveness.

In model (2), we investigate the impact of different failure rates on the spread of the epidemic in England when the vaccination rates are 0.005 and 0.01. The results shows that when the fixed vaccination rate is 0.005, the peak value when the failure rate is 0.001 is 81.4% lower than that when the failure rate is 0.01, and the peak value when the failure rate is 0.01 is 89.5% lower than that when the failure rate is 0.02. In addition, when the failure rate is 0.01, the peak time is 528 days later than when the failure rate is 0.001 and 295 days later than when the failure rate is 0.05. Therefore, the peak of epidemic situation will decrease with the decrease of failure efficiency. The peak time of the epidemic has two situations. When the failure rate is less than 0.01, the peak time will advance with the decrease of failure efficiency; when the failure rate is greater than 0.01, the peak time will be delayed with the decrease of failure efficiency. On the 60th day of vaccination, the vaccine with failure rate of 0.002 is 5.8% lower than the vaccine with failure rate of 0.01, on the 70th day of vaccination, the vaccine with failure rate of 0.002 is 9.1% lower than the vaccine with failure rate of 0.01. Therefore, with the extension of time, the vaccine with low failure rate is more effective in reducing the number of cases than the vaccine with high failure rate. When the fixed vaccination rate is 0.01, the peak of epidemic situation will decrease with the decrease of failure efficiency. When the failure rate is less than 0.019, the peak time will advance with the decrease of failure efficiency; when the failure rate is greater than 0.019, the peak time will be delayed with the decrease of failure efficiency. We also investigate the impact of different vaccination rates on the spread of the epidemic in England when the fixed vaccine failure rate is 0.005. The peak of epidemic situation decrease with the increase of vaccination rate, and the peak time is advanced with the increase of vaccination rate. When the vaccination rate is 0.025, the peak value decrease by 74.8% and the peak time is 114 days earlier than that when the vaccination rate was 0.005.

## 4 Discussion

Discussion on the rationality of the fitting value of *P* = 1.8888×10^−6^. If this fitting value refers to the decline level of antibody of actual restorers, including not only the number of officially counted inpatients, but also the number of moderate and mild patients who have not been counted, and even the number of asymptomatic infected people who have recovered by themselves, then this fitting value is obviously low. If only the recovery number of hospitalized patients is counted according to the official data of the British government, this fitting value may have practical significance. Because the antibody level of severe rehabilitation patients is relatively high, the decline of antibody level needs a longer time^[17]^.

The vaccine failure rate referred to in this paper is a vaccine failure rate in a broad sense, including the vaccine failure rate of the vaccine itself due to quality problems, including the vaccine failure rate caused by vaccination failure caused by human factors in the vaccination process, it also includes the vaccine ineffective rate who fail to produce antibodies or produce antibodies due to differences in human mechanisms, but still have the risk of infection due to low antibody levels. In practice, there may be more cases leading to vaccine failure. However, in the actual vaccination work, the vaccine failure rate can be compensated by multiple vaccination.

In the model (2) of this paper, the meaning of vaccine failure rate refers to the reciprocal of duration of vaccine effectiveness, that is, each assumed failure rate *ω* corresponds to a valid duration of vaccine effectiveness, and the data details are shown in Table 1. Similarly, each assumed vaccination rate corresponds to a time period for completing vaccination, for example,*α* = 0.005 means that it takes 1 / 0.005 days to complete all vaccination.

In the part of numerical simulation, generally speaking, the peak value always decreases with the decrease of vaccine failure rate. The peak arrival time may be related to a boundary value. When the failure rate is less than this boundary value, the peak time will advance with the decrease of failure rate; when the failure rate is greater than this boundary value, the peak time is delayed with the decrease of failure rate. In this study, when we assume that the vaccination rate is 0.005, the boundary value of failure rate is 0.01; when the vaccination rate is assumed to be 0.01, the boundary value of failure rate is 0.019. However, it is worth noting that the simulation results of the boundary value in this paper are obtained based on the failure rate taking 0.001 as the step. Strictly speaking, when we take a smaller step of failure rate for simulation, we may get a more accurate boundary value of failure rate.

Discussion on peak variation trend in simulation results. In Fig. 9 and 10, it is not difficult to find that when the failure rate increases in a fixed step, the increase of the peak is more and more obvious. The reason is that each failure rate corresponds to a duration of vaccine effectiveness. The same step of failure rate does not mean the same increase in the vaccine effective time period. In the dynamic model, we are used to using rate to study problems. In practice, we pay more attention to the duration of vaccine effectiveness.

Discussion on the significance of peak time variation trend. When the failure rate is greater than the boundary value and the peak time is delayed with the decrease of failure rate, although the epidemic peak is high (Fig.7), the delay of peak time will give the regional medical system enough time to deal with the outbreak of the epidemic. When the failure rate is less than the boundary value and the peak time is advanced with the decrease of failure rate, although the peak time comes early (Fig.6), the peak is low, so the medical system will not collapse. Therefore, no matter what the failure rate is, it is always meaningful to vaccinate. However, in practice, we prefer to use the number of cases as an indicator to measure the success of epidemic prevention and control. Therefore, it is very important to improve the effectiveness of the vaccine.

When the vaccination rate is 0.005 and the vaccine is continuously vaccinated for 70 days, the cases with failure rate of 0.002 are only 9.1% lower than those with failure rate of 0.01. The reason is that the vaccination is a continuous process, and the significant effect of the vaccine in reducing cases needs to be observed for a longer time.

Israel has one of the highest vaccination rates in the world^[18]^.On December 20, 2020, this country took the lead in launching the universal vaccination plan of COVID-19 vaccine, which is the earliest in the world. According to the data on the official website of the Israeli Ministry of health ^[19]^, as of March 2, 2020, the proportion of receiving one dose of vaccine in Israel was 51.67%, and the proportion of receiving two doses of new crown vaccine was 37.53%. By early August 2021, about 62% of Israel’s 9.3 million population had received at least one dose of vaccine, and about 58% had completed two doses of vaccine. Although Israel had reached such a high vaccination rate, it ushered in a new case peak in early September 2021. According to the data of the World Health Organization ^[5]^, on September 1, 2021, Israel added more than 10000 cases a day. Singapore is also a country with a similar situation. By the end of August 2021, the vaccination rate in Singapore had reached 80%, however, according to the data of the World Health Organization ^[5]^, since September 2021, the number of new cases in this country had increased significantly. By the end of October, more than 5000 cases had been added in a single day.

The main content of this paper is to explore the impact of different vaccine failure rates on the spread of COVID-19 epidemic in England under the condition of a certain vaccination rate. Although many countries have reached a high vaccination rate, and even reached the vaccination rate of group immunization in theory, from the data alone, vaccination does not seem to have a significant inhibitory effect on the epidemic situation in these countries. The reason may be that the mutated virus is difficult to deal with, or the effectiveness of the vaccine is not as effective as the published results. In the next article, we will use the data of severe cases to evaluate the effectiveness and protection of the vaccine. The vaccination rate can be improved through the formulation of policies by the government, however, if the protection rate of the vaccine itself is not high, increasing the vaccination rate of the vaccine will hardly help fundamentally control the epidemic. Therefore, in terms of the current situation, the evaluation of vaccine protection rate is particularly important.

## Data Availability

All data produced in the present work are contained in the manuscript.

## Acknowledgments

This study was funded by Natural Science Foundation of China (NSFC 11871093), Postgraduate Teaching Research and Quality Improvement Project of BUCEA (J2021010), BUCEA Post Graduate Innovation Project (2021098, 2021099).We thank all the individuals who generously shared their time and materials for this study.

## Notes

### Competing Interest Statement

The authors have declared no competing interest.

### Funding Statement

This study was funded by Natural Science Foundation of China (NSFC11871093), Postgraduate Teaching Research and Quality Improvement Project of BUCEA (J2021010), BUCEA Post Graduate Innovation Project (2021098, 2021099).We thank all the individuals who generously shared their time and materials for this study.

